# SARS-CoV-2 RNA detection using pooling of self-collected samples: Simple protocol may foster asymptomatic surveillance

**DOI:** 10.1101/2020.10.05.20205872

**Authors:** Giselle Ibette Lopez-Lopes, Rita de Cassia Compagnoli Carmona, Valéria Oliveira Silva, Cintia Mayumi Ahagon, Lincoln Spinazola do Prado, Fabiana Pereira dos Santos, Daniela Bernardes Borges da Silva, Katia Corrêa de Oliveira Santos, Margarete Aparecida Benega, Willian Nery Ribeiro, Audrey Cilli, Elaine Monteiro Matsuda, Ana Maria Sardinha Afonso, Maria do Carmo Sampaio Tavares Timenetsky, Luís Fernando de Macedo Brígido

## Abstract

1

**Background:** Surveillance of COVID infection and isolation of infected individuals is one of the available tools to control the spread of SAR-CoV-2. Asymptomatic and pre symptomatic are responsible for substantial transmission. RNA or antigen tests are necessary to identify non-symptomatic individuals. We tested the feasibility of using samples pooling offering different collection alternatives (swab/throat wash/saliva) to volunteers of a public health institute.

**Methods:** We evaluated pool samples from frozen material from previously tested samples and a prospective collection from asymptomatic volunteers. Some collections were paired for comparison. Pools and some individual samples were extracted with *QIAamp Viral RNA Mini Kit* (*Qiagen*, USA) and/or *Lucigen Quick Extract DNA* extraction solution (BioSearch, USA) and submitted to rtPCR (Allplex, Seegene, Korea).

**Results:** A total of 240 samples from 130 new collections and 37 samples with known result were evaluated. Pool CT was generally higher than individual samples. Lucigen extraction showed higher CT, including false negative results for samples with high CT at Qiagen extraction. Paired Swab and TW samples showed comparable results. No volunteer from negative pools reported any symptom in the 2-3 days after collection.

**Conclusions:** Clinical samples pooling to detect SARS-CoV-2 RNA is feasible and an economical way to test for COVID-19, especially in surveillance strategies targeting more infectiousness, higher viremia individuals. The use of *Lucigen* reagents show lower sensibility that may lead to false negative results with lower viremia samples. Combining throat wash with saliva may provide and interesting self-collection alternative, but more comparative work is needed.

## 2 Introduction

Sars-Cov-2 emergence from a zoonotic transmission (Gorbalenya 2020) has imposed marked social, medical and economical adaptations to the world since its description early in 2020. At the end of September, in less than a year of its recognition, over one million deaths have been associated to the COVID-19 pandemic (WHO 2020).

Although the pandemic shows signs of relative control in parts of the world, recrudescence is a constant threat, and it’s been observed in many areas (WHO 2020) specially were cases are still more substantial and surveillance suboptimal. Different vaccines products are in an advance state of development (WHO 2020b) and many show evidence of immunogenicity (reviewed by Alturki 2020), but the actual efficacy is not yet determined. Real world effectiveness will depend in different aspects that varied form general conditions, as type of cold chain needed, to more specify aspects as distinct populations response. Some crucial issues, as the durability of protection, will only be proper evaluated with time. Therefore, surveillance strategies, even in a post-vaccine era, will still be needed.

Circulation of an infected person and subsequent exposed of a susceptible individual to the virus, from exhaled air or fomites, surfaces with infectious particles from infected individuals, are the motor of the pandemic. Due to the lack of information on what constitutes immunity to infection or the durability of immunity in recovered cases (Hellestein 2020), all individuals may be considered at least partially susceptible to infection, even those with prior evidence of infection. Cumulative data since early in the epidemic have suggested that a window of about a week or two, starting 1-2 days before symptoms, a period associated to high viremia in the airways, is responsible for much of the transmission (He 2020). A major deterrent to surveillance is the fact that asymptomatic individuals, along with pre symptomatic cases, are responsible for many infections (reviewed by Huff & Singh, 2020). Association of transmission to higher viremia levels (La Scola 2020, Larremore 2020) suggests that even tests less sensible than the current real-time PCR (rt-qPCR) tests may be useful to curb transmission. The development of point of care, easy to apply tests to detect viral antigens or nucleic acid will be necessary to easy life constrains in a no-vaccine or partially effective vaccine world. Antigen tests may soon work as a surrogate to this situation. Tests that identifies infectiousness; more than viral components are urgently needed. These tests are neither available or are too costly to most of the population, so we must resort to current tools.

One of the limitations of current standard test is the need of a health care worker, with proper PPE to collect a nasopharyngeal swab. Self-swab collection, along with alternative collection strategies, as saliva and throat wash, has provided some flexibility. With the growing literature on saliva detection (Azzy 2020, Khurshid 2020, Wyllie 2020) and in throat wash efficiency for obtaining viral RNA (Saito 2020, Ali 2020), combining the methods seems adequate.

We introduced a voluntary throat wash plus saliva collection, offered to asymptomatic individuals from different areas of the institute, and tested the feasibility of using samples pooling (Abdalhamid 2020, Singh 2020, Yelin 2020) to minimize reagents and equipment use.

## 3 Methods

We evaluated pool samples both from reconstituted material from previous positive and negative tests and a prospective collection of asymptomatic volunteers.

### 3.1 Reconstituted pools

A total of 6 pools were prepared from frozen samples previously tested individually, obtained by nasopharyngeal (NP) swab or throat wash, prepared according to description below:

*1st Pool size of 10 samples: used 100 uL of each of the 10 negative samples (Lucigen), for a final volume of 1mL;*

*2nd Pool size of 10 samples: used 100 uL of each of the 03 samples of positive patients added to 100 uL of each of the 07 samples of negative patients, for a final volume of 1mL;*

*3rd Pool with size of 10 samples: used 100uL of each of the 02 positive samples added to 100uL of each of the 8 negative patient samples, for a final volume of 1mL;*

*4th Pool size of 10 samples: used 100 uL of 01 positive sample added to 100 uL of each of the 09 negative samples, for a final volume of 1mL;*

*5th Pool size of 05 samples: used 100uL of 01 positive sample added to 100uL of the first 04 samples of the 4th Pool, for a final volume of 500uL;*

*6th Pool size of 05 samples: used 100uL of 01 positive sample added to 100uL of the 04 remaining samples of the 4th Pool, for a final volume of 500uL*.

### 3.2 Prospective pools

Pools were also obtained from samples collected prospectively from volunteers, health workers from the institute, as described in **Figure 1** (flowchart).

**Figure 1.**
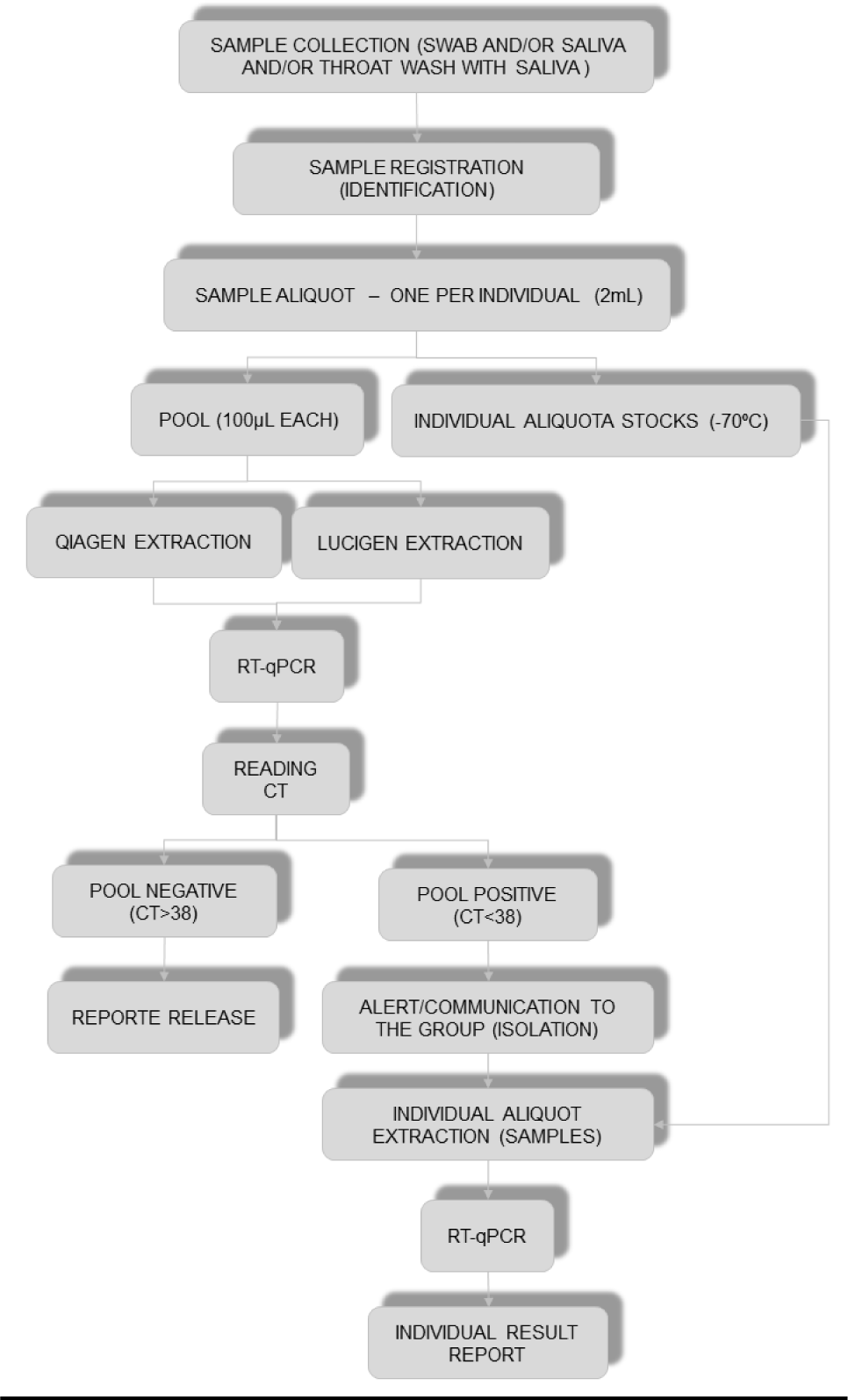
Fluxogram of prospective sample collection and pooling outcome.

Volunteers received a 5 mL cold saline (4-8 °C) in a 50 mL falcon-like tube and were oriented to perform a 5+ seconds gargle in an outside place, with safe distance from other people, followed by addition of 3-5 mL of saliva to the same tube, that was returned and kept cold or frozen (−20 °C) if not processed in the same day. Standard nasopharyngeal (NP) swab were obtained in a set of volunteers for comparison. Samples were aliquoted in a BSL-2 cabinet and pool of 3 to 12 individuals were prepared.

Pool samples were thereafter processed together with other clinical specimens following the Institute’s routine processed on the same day, with extraction using either *QIAamp Viral RNA Mini Kit* (*Qiagen*, USA) according to manufacturer instruction or *Lucigen Quick Extract DNA* extraction solution (BioSearch, USA), v/v with 40 uL of sample, followed by heating at 95 °C for 5 min and ice cooling. The sample were submitted to rt-qPCR using similar routine protocols (Allplex, Seegene, Korea), with amplification of three viral targets (E, RdRp and N) considered positive, with Human RNAse P as control. Cycle thresholds (CT) up to 37 were considered valid. In some runs, as for confirmation, the CT for the N region was the only available CT.

## 3 Results

### 3.1 Reconstituted pools

All samples selected originated from swab or throat wash collections tested in the institute with routine protocols, extracted with either *Qiagen* or *Lucigen* : 3 positive and 34 negative. Six pools were prepared, 4 of 10 samples and 2 with 5 samples. Pools were prepared with one or more positive samples with seven or more negative samples at the total of 10 samples per pool. The weaker control was further tested alone at two 5 samples pool, using different in negative samples, as shown in **Table 1** and **Table 2** shows combinations used to prepare pool.

**Table 1.** Reconstituted pool composition and number of samples

| SAMPLES CODES |  |  |  |  |  |
| --- | --- | --- | --- | --- | --- |
| *Pool 1* | *Pool 2* | *Pool 3* | *Pool 4* | *Pool 5* | *Pool 6* |
| 1 | 11 | 18 | 26 | 26 | 30 |
| 2 | 12 | 49 | 27 | 27 | 31 |
| 3 | 13 | 20 | 28 | 28 | 32 |
| 4 | 14 | 21 | 29 | 29 | 33 |
| 5 | 15 | 22 | 30 | C(+)<sup>3</sup> | C(+)<sup>3</sup> |
| 6 | 16 | 23 | 31 |  |  |
| 7 | 17 | 24 | 32 |  |  |
| 8 | C(+)<sup>1</sup> | 25 | 33 |  |  |
| 9 | C(+)<sup>2</sup> | C(+)<sup>1</sup> | 34 |  |  |
| 10 | C(+)<sup>3</sup> | C(+)<sup>2</sup> | C(+)<sup>3</sup> |  |  |

Legend: C(+) Positive controls

**Table 2.**
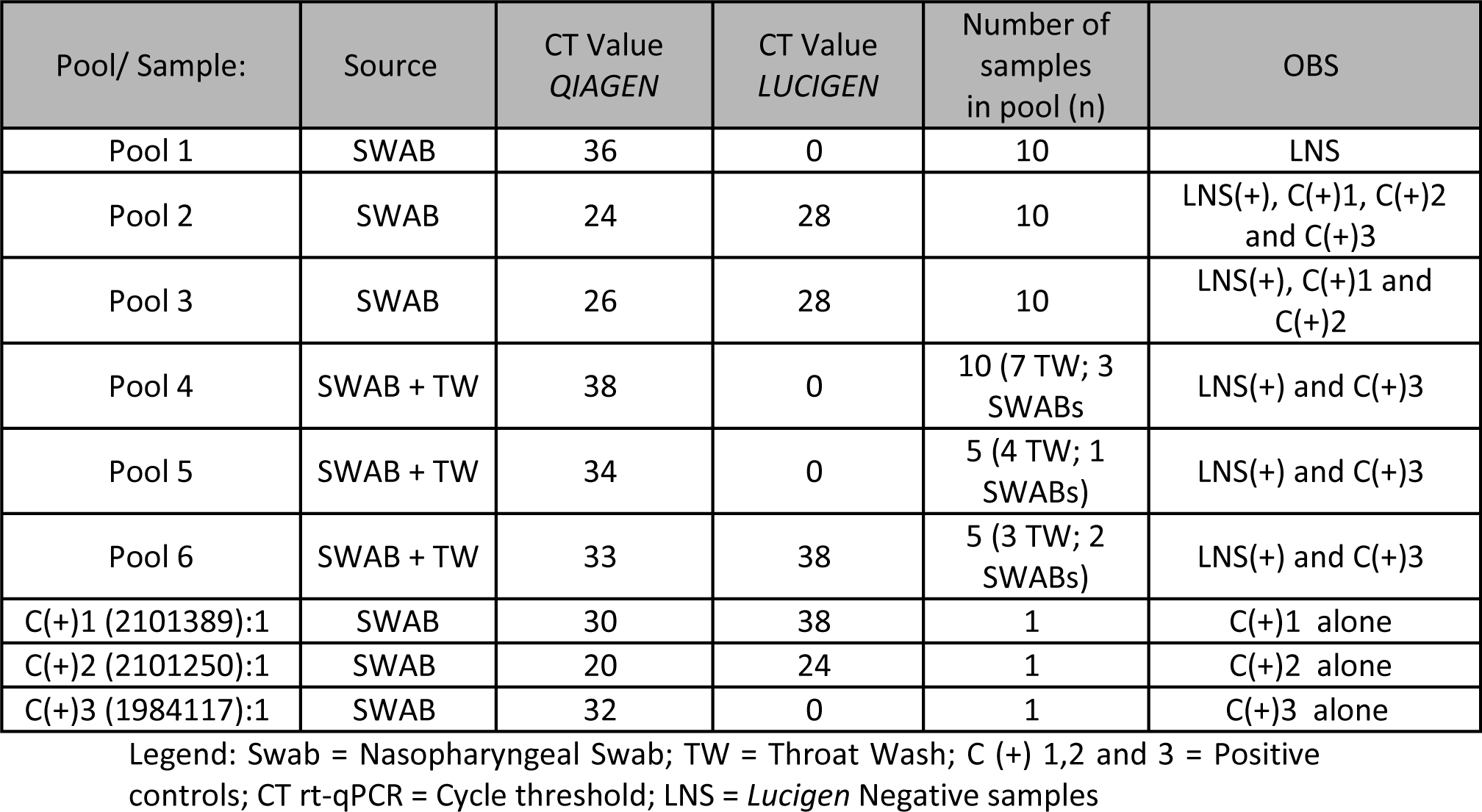
rt PCR results of reconstituted pools and individual positive control samples according to Extraction Step (Qiagen or Lucigen)

| Pool/ Sample: | Source | CT Value<br>*QIAGEN* | CT Value<br>*LUCIGEN* | Number of<br>samples<br>in pool (n) | OBS |
| --- | --- | --- | --- | --- | --- |
| Pool 1 | SWAB | 36 | 0 | 10 | LNS |
| Pool 2 | SWAB | 24 | 28 | 10 | LNS(+), C(+)<sup>1</sup>, C(+)<sup>2</sup><br>and C(+)<sup>3</sup> |
| Pool 3 | SWAB | 26 | 28 | 10 | LNS(+), C(+)<sup>1</sup> and<br>C(+)<sup>2</sup> |
| Pool 4 | SWAB + TW | 38 | 0 | 10 (7 TW; 3<br>SWABs) | LNS(+) and C(+)<sup>3</sup> |
| Pool 5 | SWAB + TW | 34 | 0 | 5 (4 TW; 1<br>SWABs) | LNS(+) and C(+)<sup>3</sup> |
| Pool 6 | SWAB + TW | 33 | 38 | 5 (3 TW; 2<br>SWABs) | LNS(+) and C(+)<sup>3</sup> |
| C(+)<sup>1</sup> (2101389):1 | SWAB | 30 | 38 | 1 | C(+)<sup>1</sup> alone |
| C(+)<sup>2</sup> (2101250):1 | SWAB | 20 | 24 | 1 | C(+)<sup>2</sup> alone |
| C(+)<sup>3</sup> (1984117):1 | SWAB | 32 | 0 | 1 | C(+)<sup>3</sup> alone |

Legend: Swab = Nasopharyngeal Swab; TW = Throat Wash; C (+) 1,2 and 3 = Positive controls; CT rt-qPCR = Cycle threshold; LNS = *Lucigen* Negative samples

Pools were tested both from *Qiagen* and *Lucigen* extracted RNA, and this results, along with individual positive control samples, are shown in table 2. Poll one, prepared as a negative control with *Lucigen* negative samples (LNS), gave a positive result in the *Qiagen* extraction procedure, the individual sample form this pool were then re-tested after *Qiagen* extraction, as described in **Table 3**.

**Table 3.**
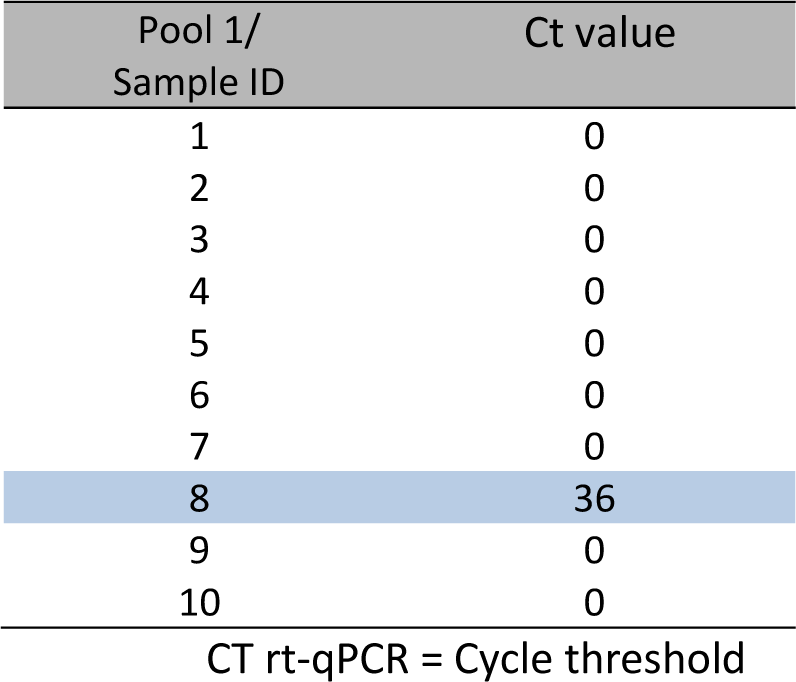
RT-qPCR SARS-CoV-19 of negative *Lucigen* samples of pool 1, tested individually after re-extraction with RNA *QIAamp Viral RNA Mini Kit* - QIAGEN.

### 3.2 Prospective pools

A total of 130 workers were tested, 83% females with a median of 47 of age (IQR 37-57), from the BioMedical, chemistry, administrative and outsourced areas. Thirty-four (26%) collected only TW, 12 (9%) collected only saliva, 22 (17%) individuals collected paired Saliva and swab samples, and 62 (48%) paired TW and swabs in a total of 214 collections from 130 volunteers. Samples were analyzed in 32 pools of 3-12 individuals each and tested using both *Qiagen* and *Lucigen* extracted RNA kits reagents.

All but two polls tested negative after *Qiagen* extraction and were not further processed. No volunteer from these pools reported any symptom in the 2-3 days after collection. Two pools from a same 6 individuals group, tested positive both in swab and TW collections. Results are shown in **Table 4**.

**Table 4.**
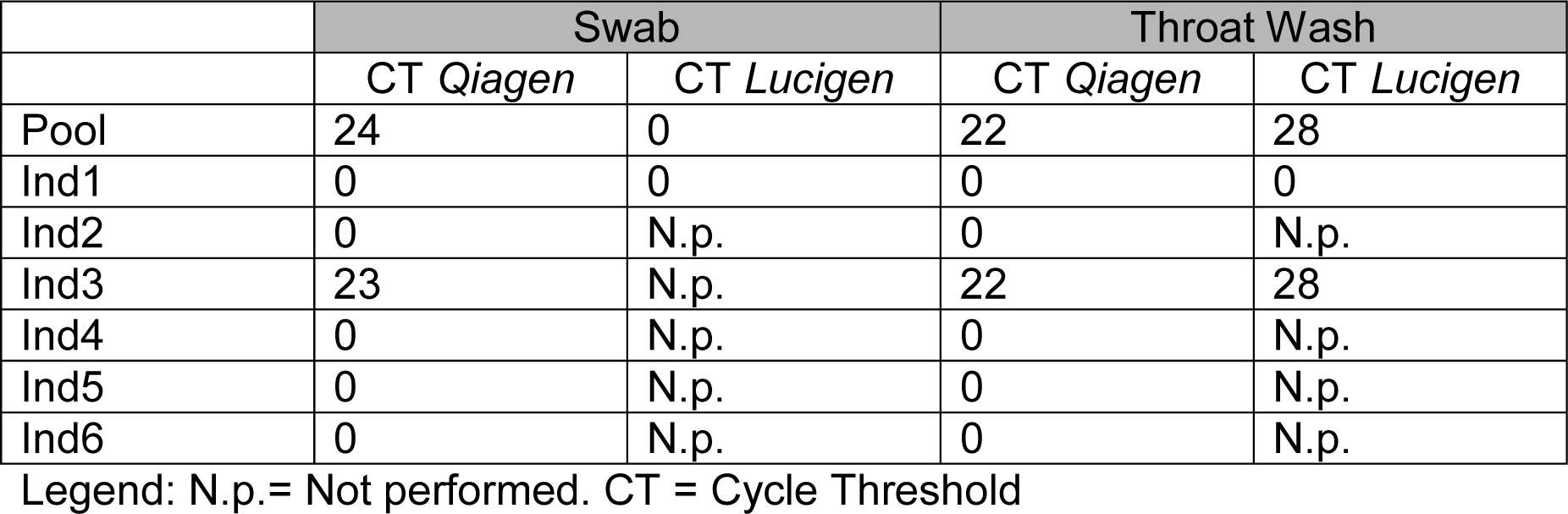
Results of positive pool per individual sample.

## 4 Discussion

In this small study, we documented the feasibility of performing an economical, easy to collect sampling of asymptomatic health care workers, analyzed in small pools (3-12) of clinical samples. Early morning collection allowed prompt processing, extraction and rt-qPCR test, providing pool results early in afternoon, with individual test of positive pool executed in an afternoon run that provided results around 7 P.M. on the same day. This swift process allowed the results to rt-qPCR individual in a short time, favoring social isolation and other pertinent attitudes.

Apart from the pooling process and contact to positive individual, the procedures involved are not different from usual routine steps, and the ability to perform the process in a swift way depends mostly in logistic and additional work load related mostly to pooling and sample identification steps. On the other hand, not only the relative high cost, especially to Low- and Middle-Income Countries (LMIC) of equipment and specialized molecular biology work force, but also the current limitations of these resources, as well as key reagents and demand for rt-qPCR machines, may suggest an important cost-effective advantage to the pooling option. We used relative small pools so we could process and give results in a same-day results, but some groups suggest larger pools, up to 32 individuals (Yelin 2020). Recently the FDA has recently stimulated the evaluation of pooling strategies (FDA 2020).

One major drawback in pooling procedure is the number of times one will need to break a pool to identify one or more positive cases. In epidemiological scenarios with high prevalence, pooling may not be attractive, but in low prevalence scenarios, as among asymptomatic monitoring in environments were infection is not yet rampant (Yates, 2020).

Pooling can provide cost effective surveillance and may actually increase testing capacity. Moreover, in some situations, pooling could be used to identify viral circulation, and the individual identification step could be omitted. This is especially true for pooling test of related people, as part of a same “epidemiological bubble”. For example, in some school returning activity, one positive test in a cohort of students may lead to home activities, for a couple of weeks, for all the group part of the pool, even in negative cases identified in individual pool partisans analysis. The current recommendation to isolate after contact to an infected person, so this may apply to all pool member when pools are constructed with interacting people..

The quarantine decision therefore may be based in the pool result itself, and does not depend on individual test. Although knowing the individual results is legitimate, in the public health perspective participants in a pool that are not involved in essential work may be quarantined when in close contact to an infected case unrestricted of the individual result.

With the lack of objective intervention for asymptomatic and mild disease, and the fact that a more severe clinical setting demands attention, even if testing negative at the sample used for the pool or tested negative individually, it may be reasonable to use pooling without individual identification in a resource limited setting, that today is not uncommon even in some wealth nations.

The characteristics of which individual should participate in a same pool in a point that may deserve attention. We tested here mostly people that had some relationship at work, as having similar activity or sharing rooms or some other kind of epidemiological link. If a pool includes only specialized people that provide essential work, it may be disruptive for the working activities to quarantine all pooling participants. In the other hand, if 4-6 workers that share some essential work activity are split in different pools, if one pool test positive, even before the individual test of the reactive pool, at least some workers may be available to perform the tasks. The pooling strategies should be customized to each epidemiological scenario and take into account the planned steps to take in case of positive pool tests.

Polling dilutes the viremia increased the CT, and the detection of low viremia cases may be compromised. Those however are not the key targets of prevention as their infectiousness is more limited (Larremore 2020, La Scola 2020). The use of *Lucigen* with heat treatment to have access to RNA for rt-qPCR, bypassing an extraction step, is simpler, cheaper and feasible, but as can be bring important increases the CT. Test for lower viremia cases, may therefore not be detected and tested as false negative without adequate extraction. In the current reagent limitations, and considering that fact that cases with lower viremia, (that is high CT) are not good transmitters (He 2020, La Scola 2020), makes its use acceptable especially in a public heath perspective, but this limitations must be taken into consideration when interpreting results from this alternative procedure.

Our work is based mostly in TW enriched with saliva. Actually, it is almost impossible guarantee a throat wash that is not “contaminated” with some saliva, and we only stimulated that more saliva is added to the TW tube. As the first step in some saliva protocols is to dilute it to make processing easier (Vaz 2020), the TW saline may act as a diluent for saliva, with the advantage to bring into the reaction more viral RNA. A combination of these methods seems interesting and TW may add to saliva based testing whenever the patient can and is willing to perform gargle and an adequate area (open air, far from others) is available. This addition is not proved yet to be relevant and saliva may perform as well. It is moreover recommended were gargle is not feasible, as for young children and at close environments.

## 5 Conclusion

Clinical samples pooling, followed by RNA extraction and routine protocols to detect SARS-CoV-2 RNA may allow increasing in the testing capability without stressing the current limitations. Although pooling may decrease sensibility, it may identify more infectious individuals and allow more frequent testing for more individuals, being a feasible, economical way to test for COVID-19 for surveillance strategies. The use of *Lucigen* reagents lead to a decrease in the sensibility that may lead to false negative results with lower viremia (higher CT) samples. Combining throat wash with saliva may increase viral recovery, but more comparative work is needed.

## Data Availability

The data presented is available for consultation

